# Immunogenicity of BNT162b2 vaccine Against the Alpha and Delta Variants in Immunocompromised Patients

**DOI:** 10.1101/2021.08.08.21261766

**Authors:** Jérome Hadjadj, Delphine Planas, Amani Ouedrani, Solene Buffier, Laure Delage, Yann Nguyen, Timothée Bruel, Marie-Claude Stolzenberg, Isabelle Staropoli, Natalia Ermak, Laure Macraigne, Caroline Morbieu, Soledad Henriquez, David Veyer, Hélène Péré, Marion Casadevall, Luc Mouthon, Frédéric Rieux-Laucat, Lucienne Chatenoud, Olivier Schwartz, Benjamin Terrier

**Author notes:** **Correspondence:** Prof. Benjamin Terrier, Département de Médecine Interne, Hôpital Cochin, 27, rue du Faubourg Saint-Jacques, 75679 Paris Cedex 14, France. These authors contributed equally to this work (Jérome Hadjadj, Delphine Planas, Amani Ouedrani). These authors contributed equally to this work (Solene Buffier, Laure Delage, Yann Nguyen, Timothée Bruel). These authors are co-senior authors (Frédéric Rieux-Laucat, Lucienne Chatenoud, Olivier Schwartz). **Collaborators (indexed in PubMed):** Philippe Blanche, Benjamin Chaigne, Pascal Cohen, Nathalie Costedoat-Chalumeau, Bertrand Dunogué, Laure Frumholtz, Gaelle Guettrot-Imbert, Véronique Le Guern, Loic Guillevin, Claire Le Jeunne, Xavier Puechal, Tali-Anne Szwebel. **Funding:**. **Declaration of interests:** T.B., I.S. and O.S. are coinventors on provisional patent no. US 63/020,063 entitled ‘S-Flow: a FACS-based assay for serological analysis of SARS-CoV-2 infection’ submitted by Institut Pasteur.

## Abstract

**Background:** The emergence of strains of SARS-CoV-2 exhibiting increase viral fitness and immune escape potential, such as the Delta variant (B.1.617.2), raises concerns in immunocompromised patients. To what extent Delta evades vaccine-induced immunity in immunocompromised individuals with systemic inflammatory diseases remains unclear.

**Methods:** We conducted a prospective study in patients with systemic inflammatory diseases (cases) and controls receiving two doses of BNT162b2. Primary end points were anti-spike antibodies levels and cross-neutralization of Alpha and Delta variants after BNT162b2 vaccine. Secondary end points were T-cell responses, breakthrough infections and safety.

**Results:** Sixty-four cases and 21 controls not previously infected with SARS-CoV-2 were analyzed. Kinetics of anti-spike IgG and IgA after BNT162b2 vaccine showed lower and delayed induction in cases, more pronounced with rituximab. Administration of two doses of BNT162b2 generated a neutralizing response against Alpha and Delta in 100% of controls, while sera from only one of rituximab-treated patients neutralized Alpha (5%) and none Delta. Other therapeutic regimens induced a partial neutralizing activity against Alpha, even lower against Delta. All controls and cases except those treated with methotrexate mounted a SARS-CoV-2 specific T-cell response. Methotrexate abrogated T-cell responses after one dose and dramatically impaired T-cell responses after 2 doses of BNT162b2.

**Conclusions:** Rituximab and methotrexate differentially impact the immunogenicity of BNT162b2, by impairing B-cell and T-cell responses, respectively. Delta fully escapes the humoral response of individuals treated with rituximab. These findings support efforts to improve BNT162b2 immunogenicity in immunocompromised individuals (Funded by the Fonds IMMUNOV; ClinicalTrials.gov number, NCT04870411).

## Introduction

The course of Coronavirus-induced disease 2019 (Covid-19) is less favorable in patients with systemic inflammatory diseases. Older age, male gender, cardiovascular disease and obesity are risk factors of severe forms and Covid-19-related death in this immunocompromised population^1–4^, as it is in the general population^5,6^. Disease-specific factors including disease activity and treatments, especially glucocorticoids, mycophenolate mofetil and rituximab, are additional risks factors^1–3^.

BNT162b2 and mRNA-1273 Covid-19 vaccines have been developed using a novel liposomal mRNA-based delivery platform. These vaccines have a good safety profile, induce strong and persistent B-cell and T-cell responses^7,8^ and are highly effective to prevent SARS-CoV-2 infection, hospitalization and death with the ancestral strain and the Alpha (B.1.1.7) variant^9^. The efficacy of vaccine has been recently mitigated by variants of SARS-CoV-2 exhibiting increase viral fitness and immune escape potential. Among them, the Delta variant (B.1.617.2) was first identified in India in October 2020 and rapidly became the predominant strain across the globe^10^. While *in vitro* data indicate reduced sensitivity of Delta variant to antibody neutralization^11^, only modest differences in vaccine effectiveness are noted with Delta as compared to Alpha^12^. In patients with systemic inflammatory diseases, the use of rituximab and methotrexate, commonly used to induce and maintain remission, decreases seroprotection rate following vaccination against influenza, pneumococcus and ancestral and Alpha variants of SARS-CoV-2^13–16^. Yet, how the different immunosuppressive or immunomodulatory drugs tune humoral and cellular responses, and how the Delta variant impacts vaccine effectiveness in this population remains unclear.

In this study, we measured seroconversion, cross-neutralization of Alpha and Delta variants and T-cell responses induced by BNT162b2 in immunocompromised patients with systemic inflammatory diseases according to the treatments received.

## Methods

### Study design

The prospective COVADIS study (NCT04870411) included patients with systemic inflammatory diseases managed in Cochin Hospital, University of Paris (Paris, France). Healthcare immunocompetent workers from the same hospital were included as controls. Patients with a positive Covid-19 serology at baseline were excluded from the main analysis. Cases and controls received two doses of BNT162b2 28 days apart. Four groups of patients receiving different immunosuppressive or immunomodulatory drugs were defined: patients receiving rituximab (“rituximab” treatment group), methotrexate (“methotrexate” group), conventional Disease-Modifying Anti-Rheumatic drugs (cDMARDs) except for methotrexate (“cDMARDs” group), and those receiving other strategies described to have limited impact on vaccine immunogenicity (“other” treatment group).

### Clinical and laboratory data

Clinical data were collected at baseline and during follow-up until month 3. To evaluate vaccine immunogenicity, blood samples were collected before the first dose of vaccine (M0), one month later just before the second dose (M1), and at 3 months (M3).

### Outcomes

Primary end points were BNT162b2 immunogenicity and cross-neutralization of Alpha and Delta variants at 3 months, i.e. after two vaccine doses, defined by neutralization titer (median of the half maximal effective dilution, ED50) for both virus with ED50 above 30. Secondary end points were the proportion of patients with positive anti-SARS-CoV-2 antibodies (define as an antibody binding unit (BU) above 1.1 for IgG and 0.2 for IgA) at M1 and M3, T-cell response defined by the number of circulating SARS-CoV-2-spike-specific IFNγ-producing T cells at M1 and M3, breakthrough infections and safety.

### T and B cell immunophenotyping

Extended B cell and T cell immunoprofiling were performed on whole blood as described in the **Supplementary Appendix** and **Supplementary Figures 1-2**.

### S-Flow assay

The S-Flow assay was used to detect antibodies bound to 293T cells stably expressing the spike protein (S) at their surface using flow cytometry. This assay is highly sensitive and allows quantification of antibodies through a standardized MFI (referred as Binding Unit; BU), which is calculated using an anti-spike monoclonal antibody as reference. The method is described in the **Supplementary Appendix**.

### Virus strains

The Alpha (B.1.1.7) variant originated from an individual returning from the United Kingdom. The Delta (B.1.617.2) variant originated from a hospitalized patient returning from India. The variant strains were isolated from nasal swabs using Vero E6 cells and amplified by two passages. Additional information is described in the **Supplementary Appendix**.

### S-Fuse neutralization assay

The S-Fuse neutralization assay was used to assess the neutralizing activity of sera against emerging variants. The method is described in the **Supplementary Appendix**.

### T-cell response using enzyme-linked immunoSpot (EliSpot)

SARS-CoV-2-specific IFNγ-producing T cells were identifed by using commercially available pools derived from a peptide scan through SARS-CoV-2 N-terminal (pool S1) and C-terminal (pool S2) fragments of spike glycoprotein (JPT Peptide Technologies GmbH, BioNTech AG, Berlin, Germany). Results are expressed as Spot Forming Unit (SFU)/10^6^ CD3+ T cells after subtracting background values from wells with non-stimulated cells. The method is described in the **Supplementary Appendix**.

### Statistical analysis

No statistical methods were used to predetermine sample size. The experiments were performed in blind regarding to the allocation groups. Flow cytometry data were analyzed with FlowJo v.10 software (TriStar). Calculations were performed using Excel 365 (Microsoft). Figures were drawn using GraphPad Prism 9. Statistical analyses were conducted using GraphPad Prism 9. Statistical significance between different groups was calculated using the tests indicated in each figure legend. Detailed statistical analysis is described in the **Supplementary Appendix**.

## Results

### Patients characteristics

Between January and April 2021, 77 cases and 28 controls were included in the study. Twenty participants (13 cases and 7 controls) with positive SARS-CoV-2 serological tests at baseline were excluded from the main analysis (**Figure 1**). Finally, 64 cases and 21 controls were analyzed. Baseline characteristics of patients are shown in **Table 1**. Median age in controls and cases was 56 [39.5-59.5] and 52 [37.8-66.3] years, respectively. The immunological characteristics are shown in **Supplementary Table 1 and Supplementary Figures 3-4**. Compared to controls, cases showed lower total lymphocytes count. As expected, in the “rituximab” treatment group, circulating B cells were not detected (except in one patient) and levels of IgG, IgA and IgM were lower.

**Table 1.** Patients’ characteristics at vaccination.

|  | All<br>n = 64 | Rituximab<br>n = 22 | Methotrexate<br>n = 16 | cDMARDs<br>n = 19 | Others<br>n = 7 |
| --- | --- | --- | --- | --- | --- |
| Age, years |  |  |  |  |  |
| Median [IQR] | 52 [37.8-66.3] | 58.5 [48.3-67.8] | 50 [38.5-72.3] | 34 [30-53.5] | 51 [44-58.5] |
| >50 yr, n (%) | 35 (54.7) | 16 (72.7) | 8 (50) | 7 (36.8) | 4 (57) |
| Female, n (%) | 48 (75) | 15 (68.2) | 11 (68.8) | 15 (79) | 7 (100) |
| **Diagnosis** |  |  |  |  |  |
| Vasculitis |  |  |  |  |  |
| AAV | 18 (28.1) | 18 (81.8) | 0 | 0 | 0 |
| Behçet's | 2 (1.6) | 0 | 0 | 2 (10.5) | 0 |
| Cryoglobulinemia vasculitis | 2 (1.6) | 2 (9.1) | 0 | 0 | 0 |
| Large vessel vasculitis | 4 (6.3) | 0 | 4 (25) | 0 | 0 |
| Connective Tissue Disease |  |  |  |  |  |
| SLE | 15 (23.4) | 0 | 4 (25) | 9 (47.4) | 2 (28.6) |
| Systemic sclerosis | 7 (10.9) | 1 (4.5) | 0 | 4 (21.1) | 2 (28.6) |
| Sjogren Syndrome | 2 (1.6) | 1 (4.5) | 1 (6.3) | 0 | 0 |
| Myositis | 5 (7.8) | 0 | 3 (18.8) | 2 (10.5) | 0 |
| Inflammatory Rheumatic Diseases* | 3 (4.7) | 0 | 2 (12.5) | 0 | 1 (14.3) |
| Sarcoidosis | 3 (4.7) | 0 | 1 (6.3) | 1 (5.3) | 1 (14.3) |
| Others | 3 (4.7) | 0 | 1 (6.3) | 1 (5.3) | 1 (14.3) |
| Disease duration (years). mean (SD) | 9.5 (9) | 9.2 (9.1) | 10.1 (8) | 8.4 (8.9) | 12 (11.9) |
| Disease activity status |  |  |  |  |  |
| Active disease n (%) | 17 (26.5) | 4 (18.2) | 6 (37.5) | 7 (36.8) | 0 (0) |
| Renal involvement n (%) | 19 (29.7) | 9 (41) | 3 (18.8) | 6 (31.6) | 1 (14.3) |
| **Ongoing treatments n (%)** |  |  |  |  |  |
| Prednisone | 45 (70.3) | 13 (59.1) | 12 (75) | 17 (89.5) | 3 (42.9) |
| Median mg/day (IQR) | 7.5 (5-15) | 5 (5-13.8) | 7.5 (5-13.8) | 10 (5-25) | 5 (5-12.5) |
| cDMARDs |  |  |  |  |  |
| Methotrexate | 19 (29.7) | 3 (13.6) | 16 (100) | 0 | 0 |
| Azathioprine | 5 (7.8) | 0 | 0 | 5 (26.3) | 0 |
| Mycophenolate Mofetil | 12 (18.8) | 0 | 0 | 12 (63.2) | 0 |
| Cyclophosphamide | 3 (4.7) | 1 (4.5) | 0 | 2 (10.5) | 0 |
| Biological therapies |  |  |  |  |  |
| Anti-TNF-α | 6 (9.4) | 0 | 1 (6.3) | 3 (15.8) | 2 (28.6) |
| Rituximab | 22 (34.4) | 22 (100) | 0 | 0 | 0 |
| Tocilizumab | 3 (4.7) | 0 | 3 (18.8) | 0 | 0 |
| Belimumab | 1 (1.6) | 0 | 1 (6.3) | 0 | 0 |
| Hydroxychloroquine | 15 (23.4) | 2 (9.1) | 4 (25) | 7 (36.8) | 2 (28.6) |
| No DMARDs, biologics or prednisone | 1 (1.6) | - | - | - | 1 (14.3) |
| **Number of lines of previous treatments. n.**<br>**median (IQR)** | 2 (1-3.8) | 2 (1-4.3) | 2 (1-4) | 2 (1-3) | 2 (1-2) |

\* Inflammatory Rheumatic Diseases : rheumatoid arthritis (n=2), spondyloarthritis (n=1).

**Figure 1.**
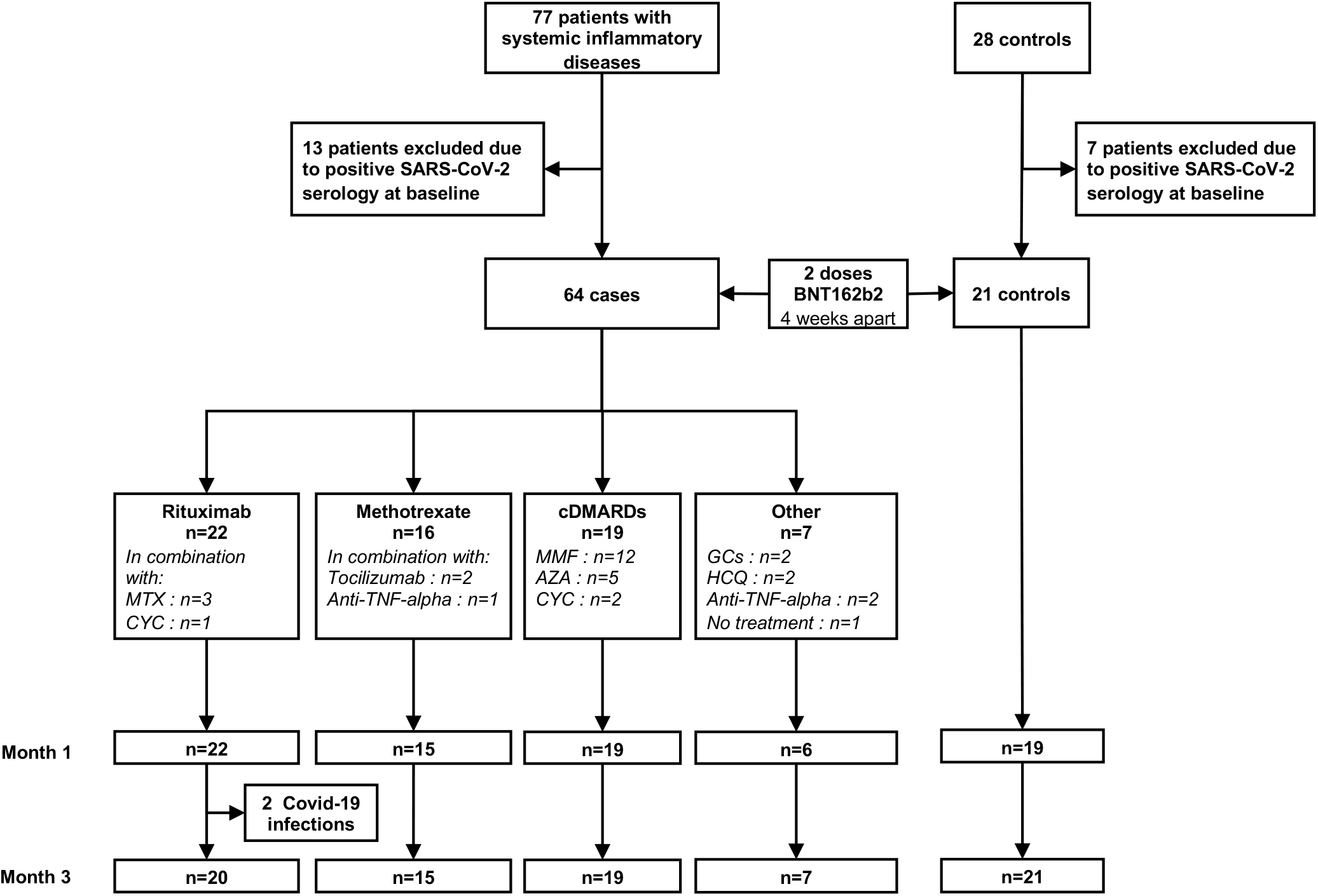
Flow-chart of the study.

### Induction of anti-spike antibodies after BNT162b2 vaccine

First, we analyzed the kinetics of induction of anti-spike IgG and IgA in patients’ sera after the first and second dose of BNT162b2 vaccine. We observed a delayed response in cases compared to controls. Anti-spike IgG and IgA inductions were detectable mainly after the second dose in cases, whereas it was noted from the first dose in controls (**Supplementary Figure 5**). On samples collected after the 2 doses, at 3 months, all treatment groups except the “other” group showed significantly lower anti-spike IgG levels than controls (**Figure 2A**). The “rituximab” group showed the lowest response. Then, we categorized individuals who seroconvert in IgG or IgA at M3 as “responders”. All controls and cases from the “other” treatment group seroconverted in IgG (**Figure 2B**). “Rituximab” showed again the lowest response, with only 50% of individuals who seroconverted at M3 (**Figure 2B)**. “Methotrexate” and “cDMARDs” treatment groups showed intermediate levels of anti-spike IgG levels at 3 months, with 93% and 68% of individuals who seroconverted, respectively (**Figure 2A-B)**. A large inter-individuals variability was observed in these two groups. The use of azathioprine or mycophenolate mofetil did not discriminate between responders and non-responders in the cDMARDs group. Anti-spike IgA levels after two doses of BNT162b2 were lower than for IgG in all cases and controls (**Figure 2A-B**). Anti-spike IgA levels after the first and second doses of BNT162b2 vaccine were significantly lower in patients treated with rituximab and methotrexate compared to controls and cases from the “cDMARDS” and “other” treatment groups (**Figure 2A and 2B** and **Supplementary Figure 5**). Analysis of the circulating follicular helper CD4+ T cells after the first and second dose showed a delayed increase in cases compared to controls, occurring mainly after the second dose in patients treated by methotrexate and cDMARDS and detected after the first dose in controls. No difference was observed in the proportion of memory B cells (**Supplementary Figure 6)**.

**Figure 2.**
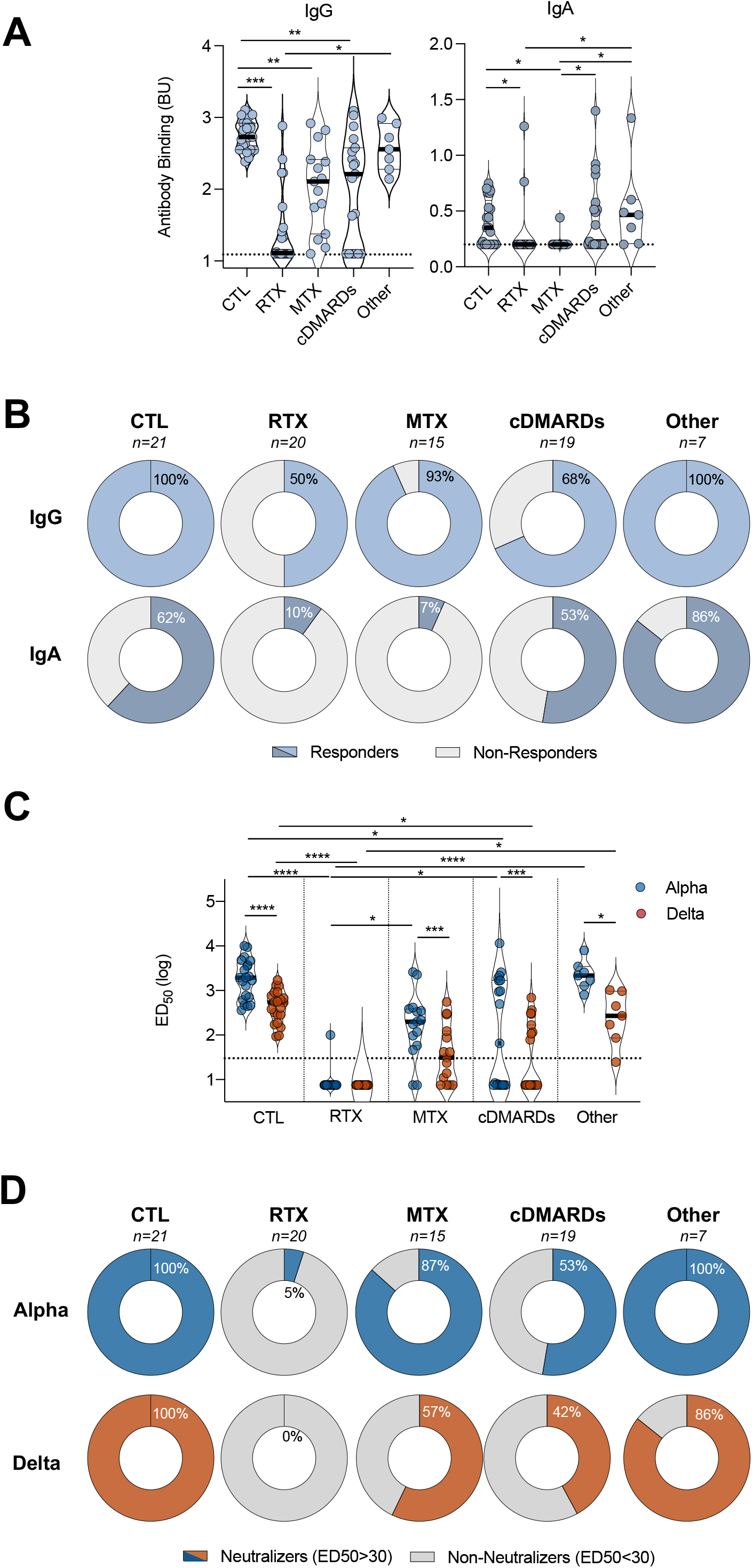
Humoral Immune Response to SARS-CoV-2 3 Months After BNT162b2 Vaccine. **A**. Levels of anti-S IgG and IgA antibodies in the indicated groups after full vaccination at 3 months (M3) as determined by the SFlow assay. The Binding Unit (BU), in a log scale, is calculated using a serially diluted anti-S monoclonal antibody as standard. Dotted lines indicate threshold of positivity (BU=1.1 for IgG and 0.2 for IgA). Two-sided Kruskal-Wallis test with Dunn’s test for multiple comparisons were performed. *P<0.05, **P<0.01, ***P<0.001, ****P<0.0001. **B**. In each group, individuals were defined as a ‘responders’ (blue) if antibodies were detected above the threshold or non-responders’ (gray) otherwise. Numbers of individuals in each group and percentages of responders are indicated. **C**. Neutralizing titers of sera against Alpha and Delta variants are expressed as ED50 values, in a log scale. Dotted line indicates the limit of detection (ED50=30). Data are mean of two independent experiments. In each group, Wilcoxon paired t-test was performed to compared ED50 of Alpha *versus* Delta variants. Two-sided Kruskal-Wallis test with Dunn’s test for multiple comparisons between group of treatment was performed. *P<0.05, **P<0.01, ***P<0.001, ****P<0.0001. **D**. In each group, individuals were defined as a ‘neutralizers’ (blue for Alpha; orange for Delta) if neutralization was detected at the dilution 1:30 or ‘non-neutralizers’ (gray) otherwise. Numbers of individuals in each group and percentages of neutralizers are indicated.

### Neutralization of Alpha and Delta variants by sera after BNT162b2 vaccine

We next examined whether BNT162b2 vaccine-elicited antibodies at month 3 neutralized the Alpha and Delta variants in cases and controls (**Figure 2C-D**). Median ED50 for Alpha in controls and in cases from the “rituximab”, “methotrexate”, “cDMARDs” and “other” treatment groups were 1942, <7.5, 199, 65 and 2173, respectively; and 539, <7.5, 31, <7.5 and 270 for Delta (**Figure 2C**). Delta was 4-fold less sensitive to neutralization than Alpha in the controls, confirming previous observation^11^. Among cases, titers were reduced by 6-fold between Delta and Alpha in the “methotrexate” group, 9-fold in the “cDMARDs” group, 8-fold in the “other” group. The lack of neutralization in the “rituximab” group impaired the calculation of a fold decrease.

Then, we arbitrarily classified individuals as neutralizers according to the detection of neutralizing antibodies at a serum dilution of 1:30 and non-neutralizers. Administration of two doses of BNT162b2 generated a neutralizing response against the Alpha and Delta variants in 100% of controls. Only one individual in the “rituximab” group neutralized Alpha (5%) and none neutralized Delta (**Figure 2C-D**). Sera of 87% of patients in the “methotrexate” group neutralized Alpha, dropping to 57% against Delta (**Figure 2D**). Sera from patients in the “cDMARDs” group neutralized Alpha and Delta in 53% and 42%, respectively. Nine (14%) cases neutralized Alpha but not Delta, including 5 patients treated with methotrexate, 2 with cDMARDs, 1 with rituximab and 1 with anti-TNF-α therapy. Correlation between Alpha and Delta neutralization titers, and between IgG production and ED50 of Alpha variant was strong in all participants except for those receiving rituximab and cDMARDs (**Supplementary Figure 7**).

The lack of neutralization of Delta was associated with active disease (P<0.001), the use of rituximab (P<0.001), glucocorticoids (P=0.007) and low IgM (P=0.047) and IgG2 (P=0.05) levels (**Supplementary Table 3**). In multivariate analysis, ED50 of Delta remained negatively associated with rituximab (P<0.001), methotrexate (P<0.001) and cDMARDs (P<0.001) (**Supplementary Table 4**).

Overall, B-cell response to BNT162b2 vaccine was impaired in immunocompromised patients at different levels depending on the treatments received. The effect was further amplified when evaluating the efficacy of sera to neutralize the Delta variant.

### Seroconversion and neutralization of Alpha and Delta variants in Convalescent Vaccinated Individuals

We then quantified anti-spike IgG and IgA and neutralization activity 3 months after vaccination in the 7 controls and 13 cases who had been previously infected with SARS-CoV-2 and excluded from the main analysis (**Supplementary Figure 8**). In convalescent controls, vaccination boosted levels of anti-spike IgG and IgA as well as neutralizing antibody titres against both variants, as compared to the uninfected vaccinated control group. In previously infected cases under immunosuppressive or immunomodulatory drugs, a low response remained after vaccination.

### T-cell Response to BNT162b2 Vaccine

We next investigated whether controls and cases mounted a SARS-CoV-2 specific T-cell response following the first and second doses of BNT162b2 vaccine (**Figure 3** and **Supplementary Figure 9**). All controls (excepte one) and cases except those from the “methotrexate” treatment group had similar levels of specific T-cells in response to S1 pool (**Figure 3A)**. Methotrexate completely abrogated T-cell responses after one dose and dramatically impaired T-cell responses after 2 doses of BNT162b2 compared to controls and cases from other treatment groups (**Figure 3A and 3B)**. Similar results, but less pronounced, were observed for S2 peptide pool (**Figure 3A and Supplementary Figure 9**). Importantly, despite the absence of neutralizing activity in response to BNT162b2, patients receiving rituximab showed increased levels of specific T-cell responses that reached after a delay the same levels as controls (**Figure 3A and 3B)**. The relationship between humoral and cellular immune responses against SARS-CoV-2 is shown in **Figure 4**, highlighting the impact of the different treatment groups on both humoral and cellular responses. The lack of T-cell response was associated with the use of methotrexate (P=0.045) and glucocorticoids (P=0.012). In multivariate analysis, no variable correlated with SARS-CoV-2-specific IFNγ-producing T cells. Also, no significant differences in the proportion of circulating CD4+ memory T cells and Th1 T cells after the first and second dose of BNT162b2 were found in all groups (**Supplementary Figure 6**).

**Figure 3.**
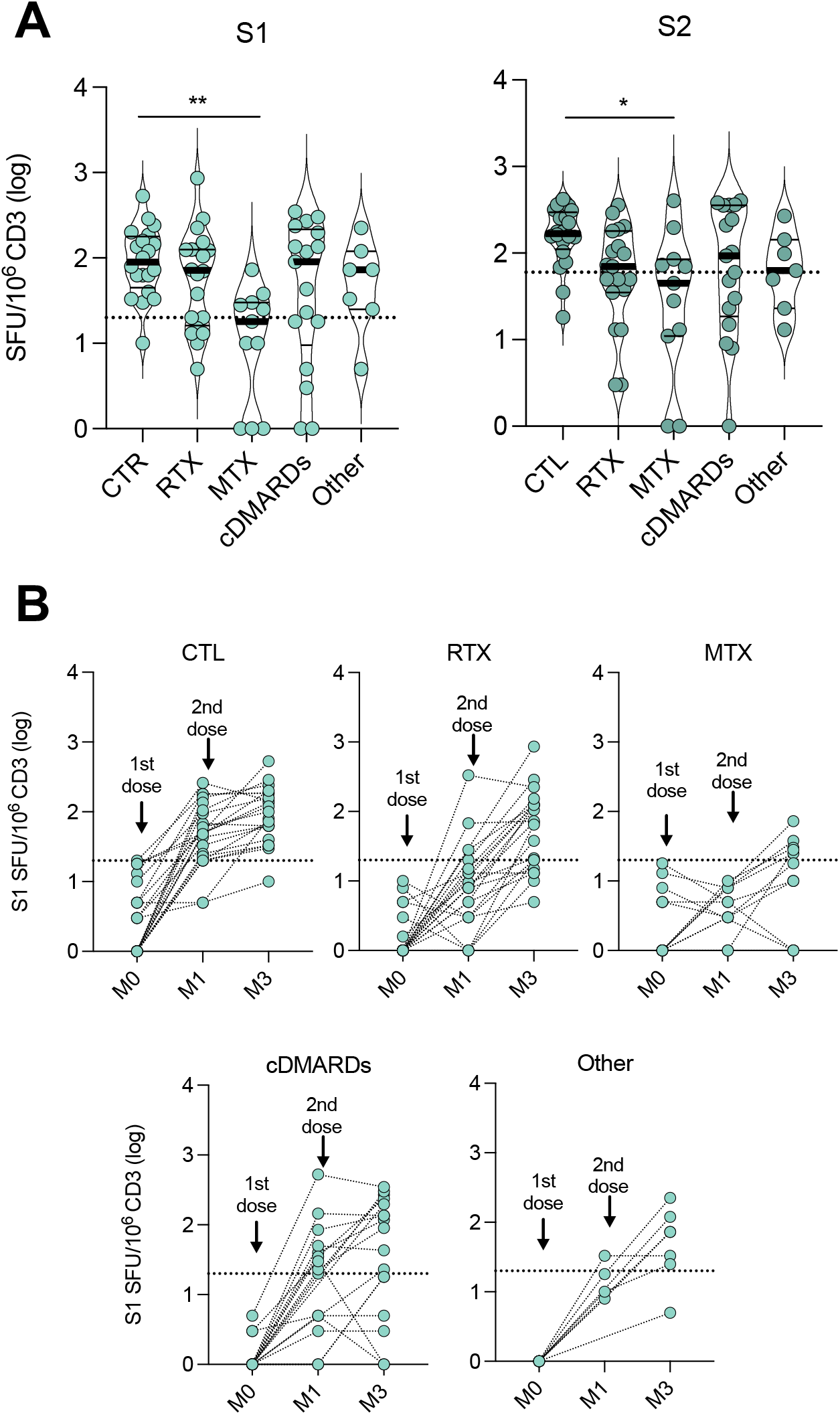
Cellular Immune Response to SARS-CoV-2 After BNT162b2 Vaccine. **A**. Quantification of SARS-CoV-2 specific T-cells responses using EliSpot at M3 in the indicated groups. Results were expressed as spot forming unit (SFU)/10^6^ CD3+ T-cells after subtraction of background values from wells with non-stimulated cells, in a log scale. Negative controls were PBMC in culture medium. Positive controls were PHA-P and CEFX Ultra SuperStim Pool. SARS-Cov-2 peptide pools tested were derived from a peptide scan through SARS-CoV-2 Spike glycoprotein (Left S1, N-terminal fragment, right: S2, C-terminal fragment). P values were determined with two-sided Kruskal-Wallis test with Dunn’s test for multiple comparisons were performed. *P < 0.05; **P < 0.01; ***P < 0.001. **B**. Kinetic of specific T-cells responses against the SARS-CoV-2 S1 peptide before the first dose (M0), before the second dose (M1) and after full vaccination at 3 months (M3) according to the treatments received. Data indicate median. Each dot represents a single patient.

**Figure 4.**
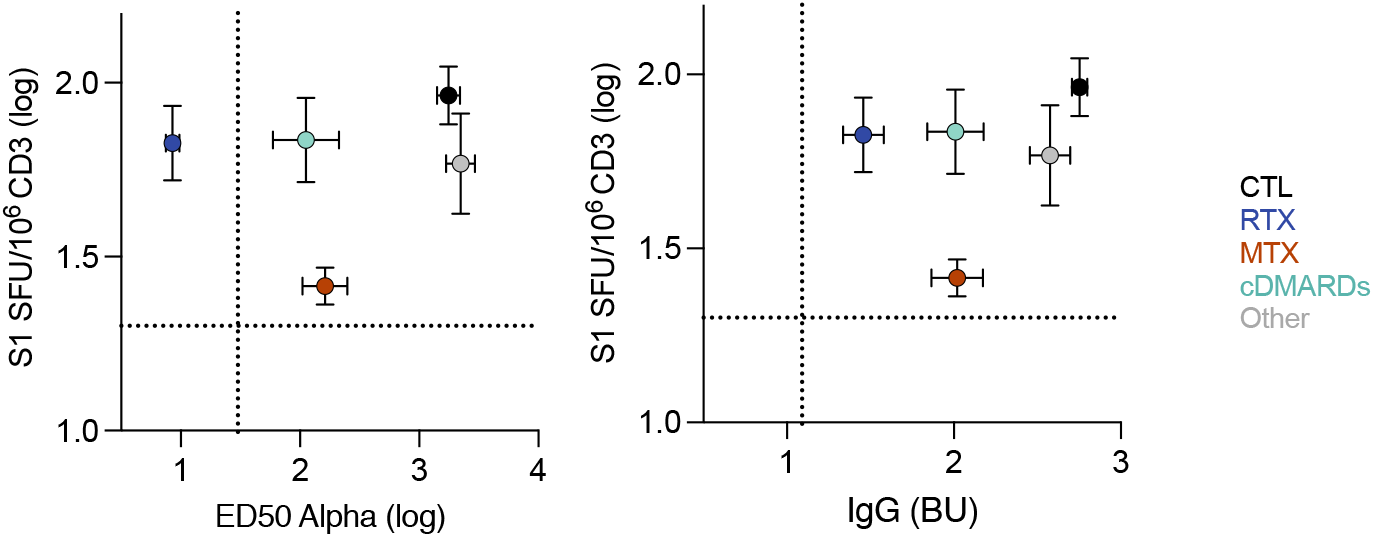
Relationship Between Humoral and Cellular Immune Responses Against SARS-CoV-2. Each dots represent the mean of all patients in the indicated groups. Bars are the standard deviation. X axis is the neutralization titer (ED50) for Alpha (left panel) or the levels of IgG (right panel). Y axis is the levels of S1-specific T-cell responses measured by ELISPOT. Dotted lines indicate the limit of detection.

Overall, T-cell responses to S1 and S2 peptide pools were similar in cases compared to the controls except methotrexate treated patients showing significantly decreased T-cell responses.

### Clinical Outcomes After BNT162b2 Vaccine

At 3 months of follow-up, 2 patients from the cohort developed Covid-19, as confirmed by a positive SARS-CoV-2 RT-qPCR test in nasopharyngeal swab for Alpha and Beta variants. Both individuals belonged to the “rituximab” treatment group. Symptoms started 4 and 7 days after the second dose of vaccine. They were treated with anti-SARS-CoV-2 monoclonal antibodies with a good outcome. Four patients (6.3%) experienced a disease flare within the 3 months following the first dose of vaccine, two patients with systemic lupus erythematosus and two with systemic vasculitis, leading to modification of immunosuppressive regimen.

## Discussion

As the Delta variant spreads across the globe, aggregating data on the effectiveness of Covid-19 vaccines in specific immunocompromised populations is a critical issue. Data from solid organ transplant recipients, patients with malignant hemopathy or with chronic inflammatory arthritis suggested that risk factors for reduced SARS-CoV-2 vaccine immunogenicity included older age and treatments with glucocorticoids, rituximab, mycophenolate mofetil and abatacept^15,17–19^. However, levels of anti-spike antibodies were mainly measured and few studies used neutralization assay or assessed T-cell response.

Additional studies specifically reported that B cell depletion by rituximab blocked humoral but not T cell response to vaccination, using anti-RBD IgG measurement and IFNγ ELISPOT T-cell response. SARS-CoV-2 antibody response was reported in 0 to 39% of the vaccinated B-cell-depleted patients, whereas T cell responses were noted in 58 to 100%^20,21^. This early assessment showed that humoral immunity to one or two doses of BNT162b2 was also impaired by methotrexate treatment^22–24^. However, conflicting results were found for cellular responses showing either preserved^22^ or impaired T-cell activation^24^. Most of these studies assessed very early timepoints that may not allow an appropriate assessment of immune response after complete vaccination.

Sera from convalescent and vaccinated individuals neutralize less efficiently the Delta variant than the Alpha^11^. However, this was studied in the general population and assessing the sensitivity of the Delta variant to antibody neutralization in immunocomprised populations is thus necessary.

In this study, we focused on patients with systemic inflammatory diseases that were receiving rituximab, methotrexate and/or other cDMARDs, and provided important data regarding sensitivity to Delta variant according to the treatments used. We analyzed patients after the first and the second doses of the BNT162b2 vaccine. We report a delayed and lower induction of anti-spike IgG and IgA compared to controls, much more pronounced with rituximab. While two doses of BNT162b2 generated a neutralizing response against Alpha and Delta variants in 100% of controls, 95% of sera from patients treated with rituximab did not neutralize these two variants. In contrast, SARS-CoV-2 specific T-cell response were similarly measured in controls and cases with the exception of methotrexate-treated patients. This differential impairement of immunogenicity after BNT162b2 vaccine according to the treatments received, mainly for rituximab and methotrexate, is critical to identify patients in which optimization of vaccine strategies should be evaluated.

To counteract this impaired immunogenicity, the administration of a third dose of mRNA-based vaccine has been proposed. Recent data in solid-organ transplant recipients showed that a third dose of BNT162b2 vaccine increased the prevalence of seroconversion and antibody titers, without serious adverse events^25–27^. A third dose also increased specific cellular response even in patients who remained seronegative^27^.

Our study has several limitations. The findings are observational and based on small numbers and should be interpreted with caution. Differences in treatment groups where highly associated with the type of underlying inflammatory disease, and there may be differences among the populations. However, except for more frequent renal involvement at diagnosis in the “rituximab” group and younger age in the “cDMARDs” group, patients’ characteristics were comparable between treatment groups.

Overall, we found that rituximab and methotrexate differentially impact the immunogenicicty of BNT162b2 vaccine, by imparing B-cell and T-cell responses, respectively. The Delta variant fully escapes the sub-optimal humoral response of individuals treated with rituximab. Our findings support efforts to improve effectiveness of mRNA vaccines in this immunocompromised population.

## Data Availability

All data are available by sending email to Pf Benjamin Terrier

## Figure legends

**Figure S1.**
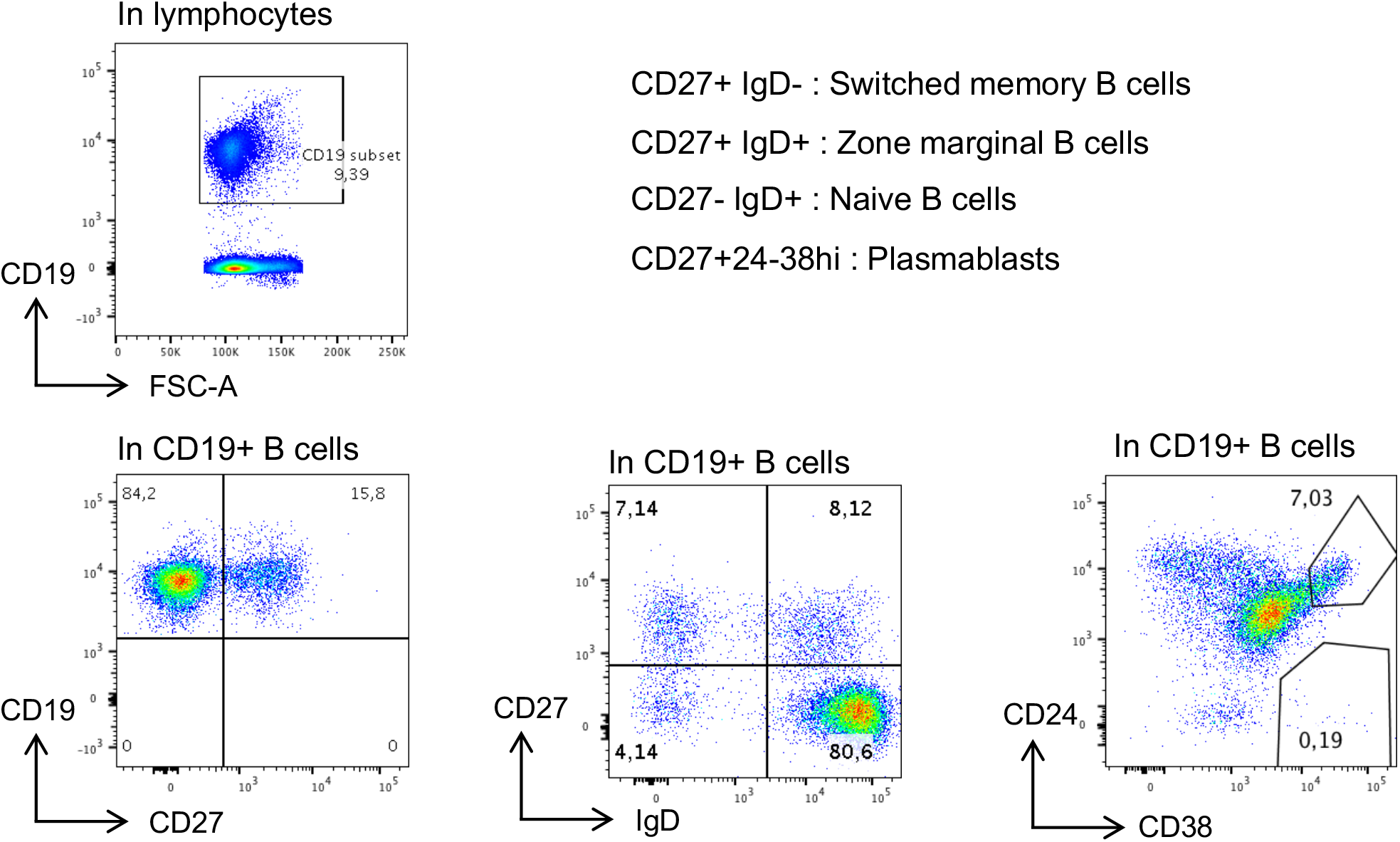
Gating strategy for immunoprofiling of B cell subpopulation compositions.

**Figure S2.**
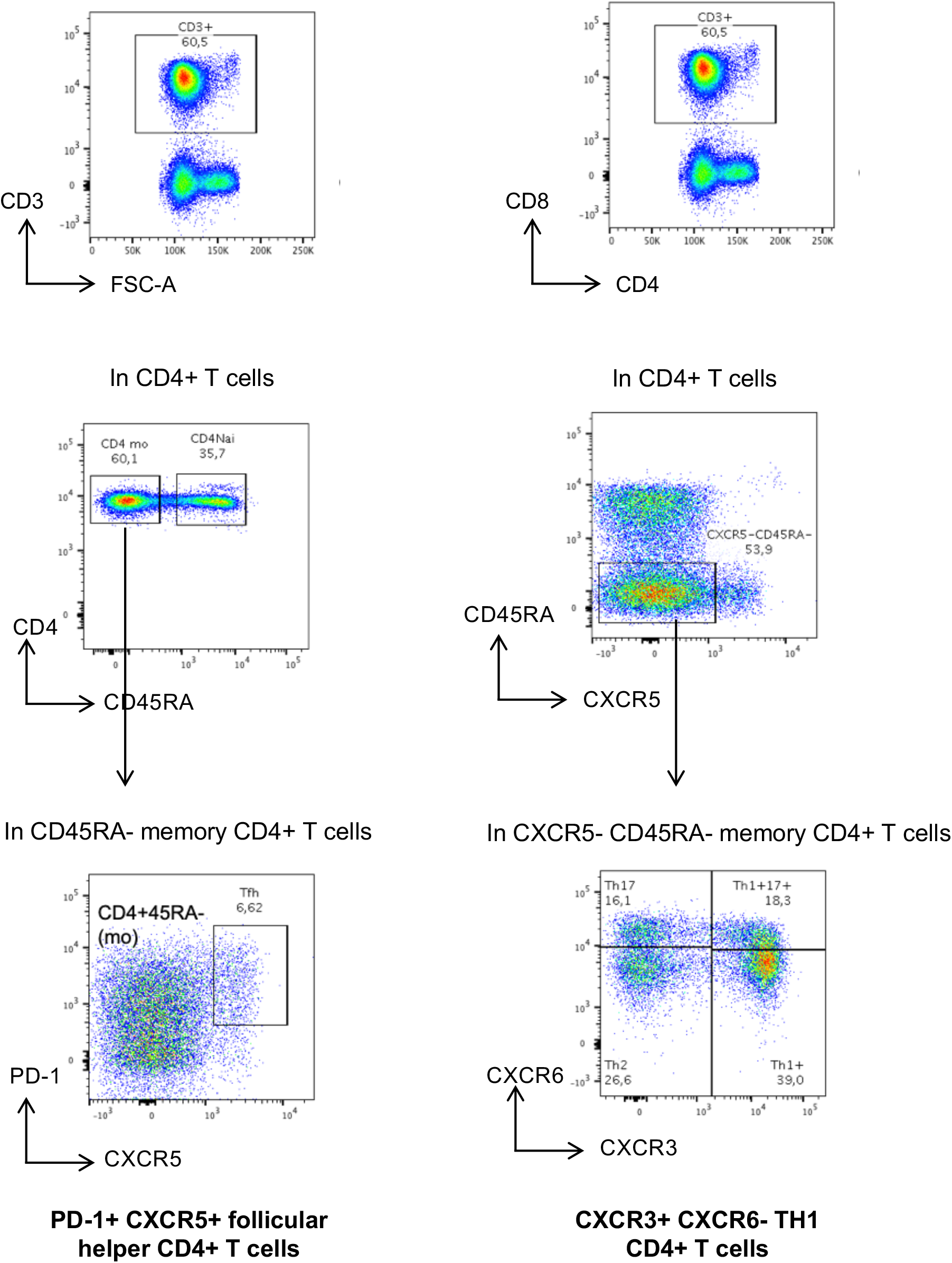
Gating strategy for immunoprofiling of T cell subpopulation compositions.

**Figure S3.**
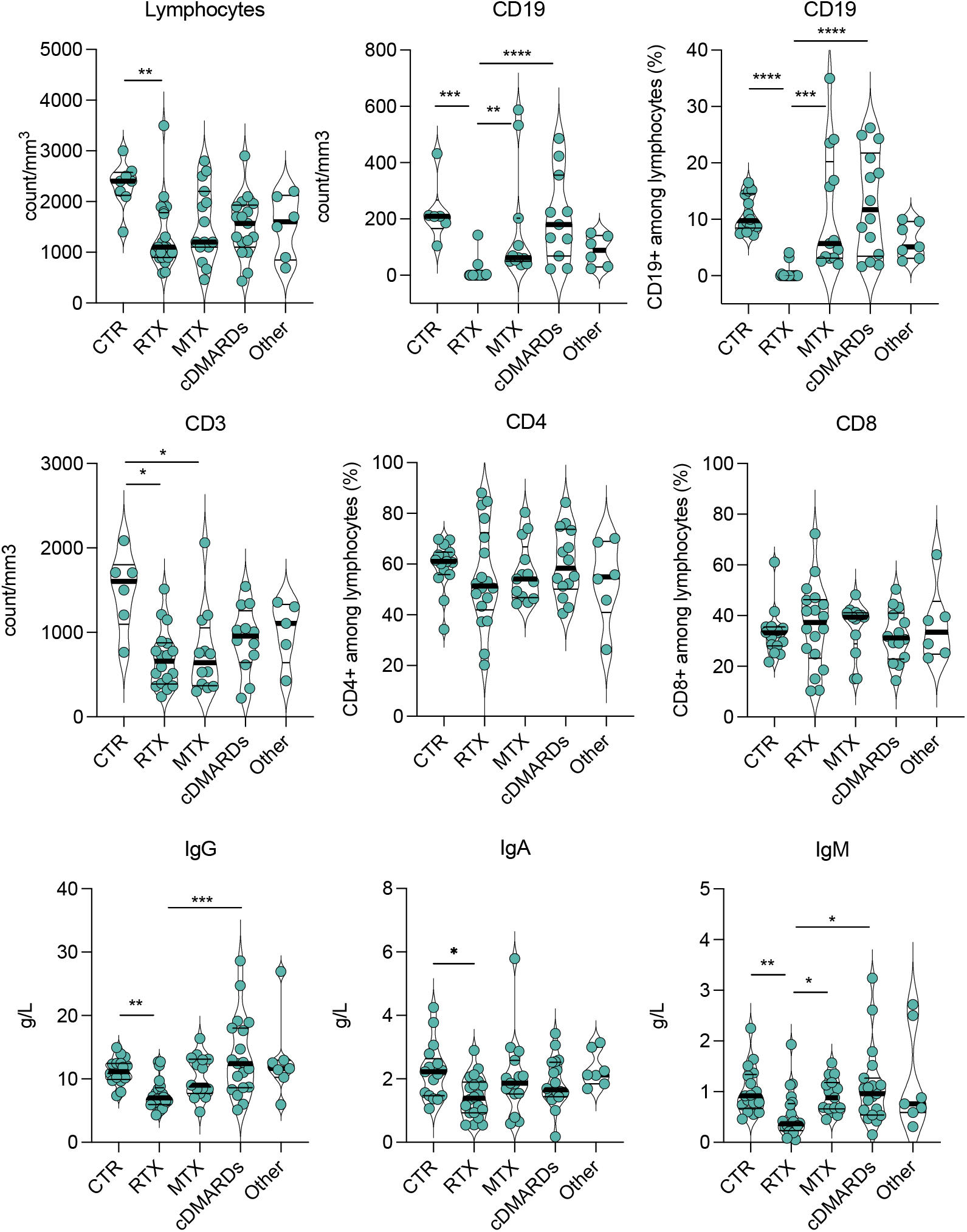
Laboratory Findings in Patients Included in the Study.

**Figure S4.**
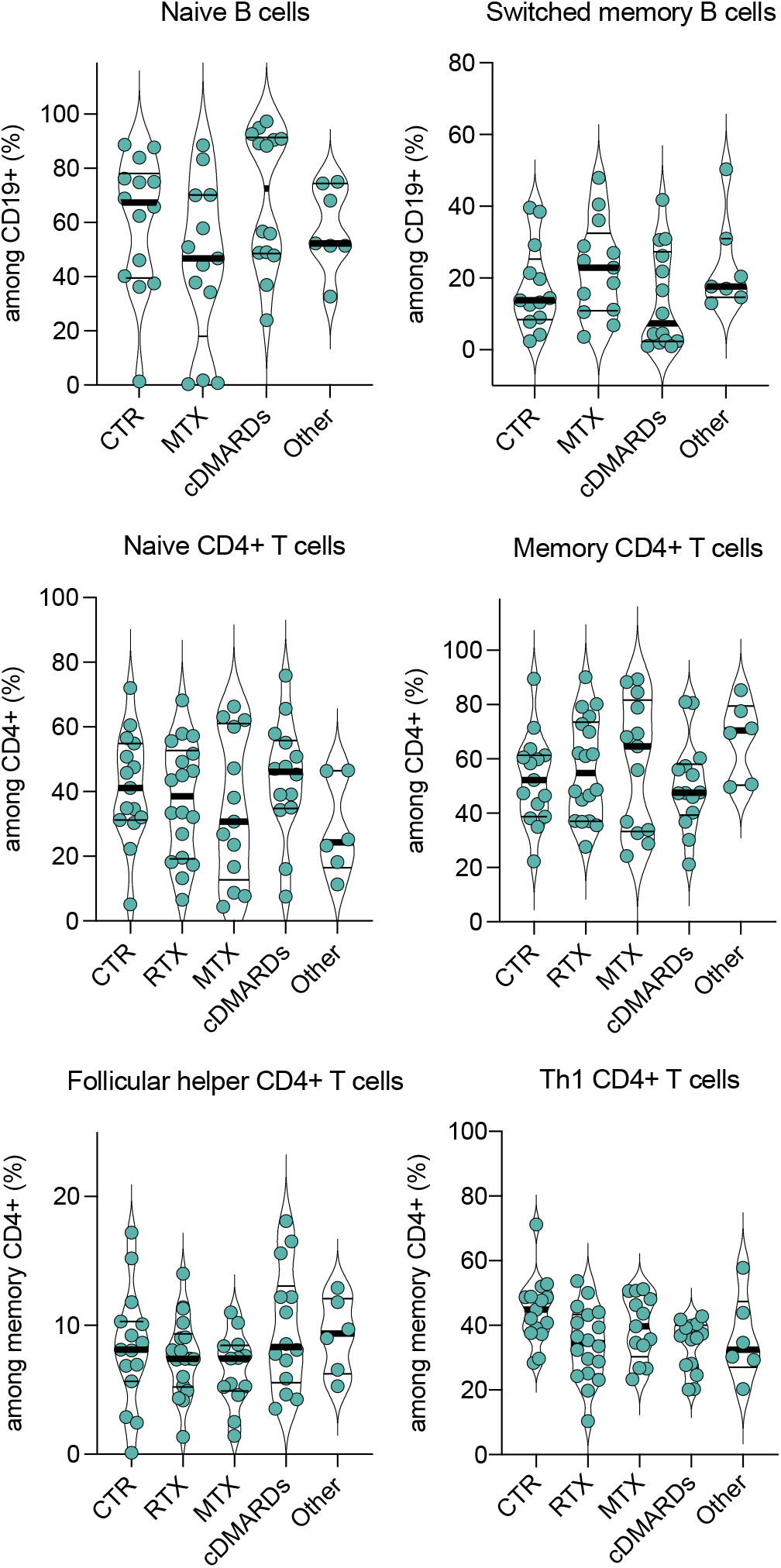
Immunoprofiling of Peripheral Blood Mononuclear Cells at Baseline.

**Figure S5.**
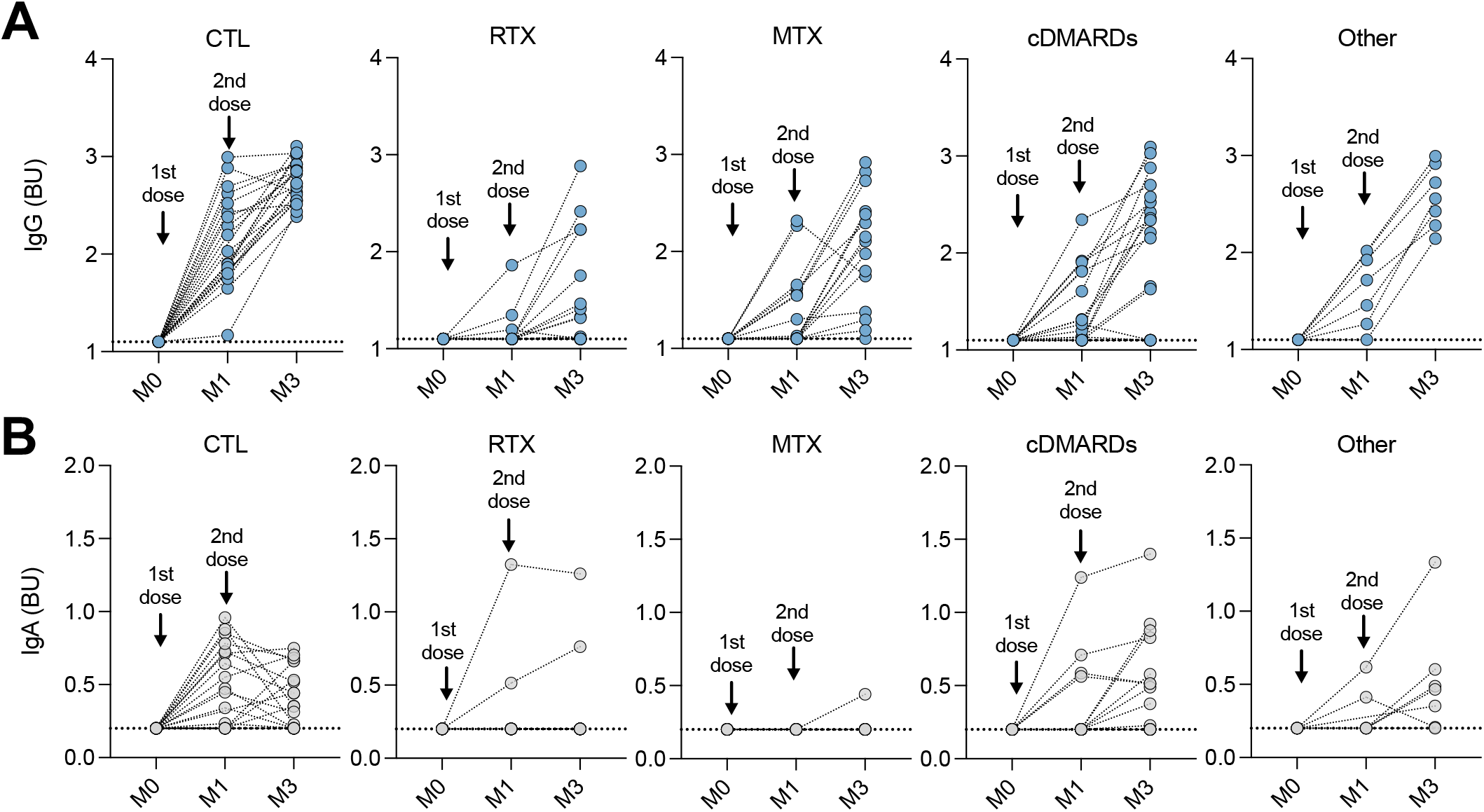
Kinetics of Anti-Spike IgG and IgA after BNT162b2 Vaccine.

**Figure S6.**
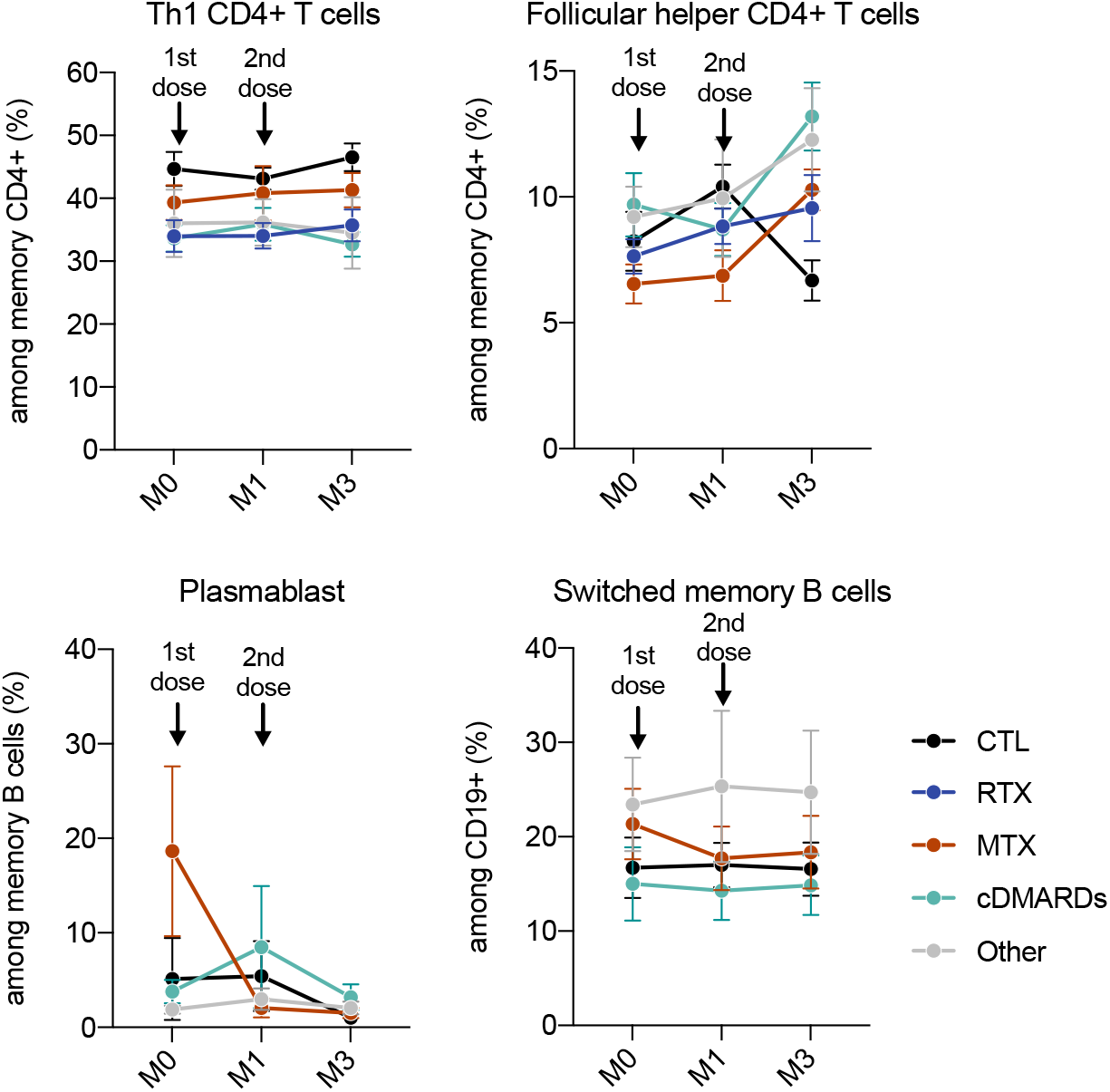
Kinetics of TH1 CD4+ T cells, Follicular Helper CD4+ T cells, Plasmablasts and Switched Memory B cells After BNT162b2 Vaccine.

**Figure S7.**
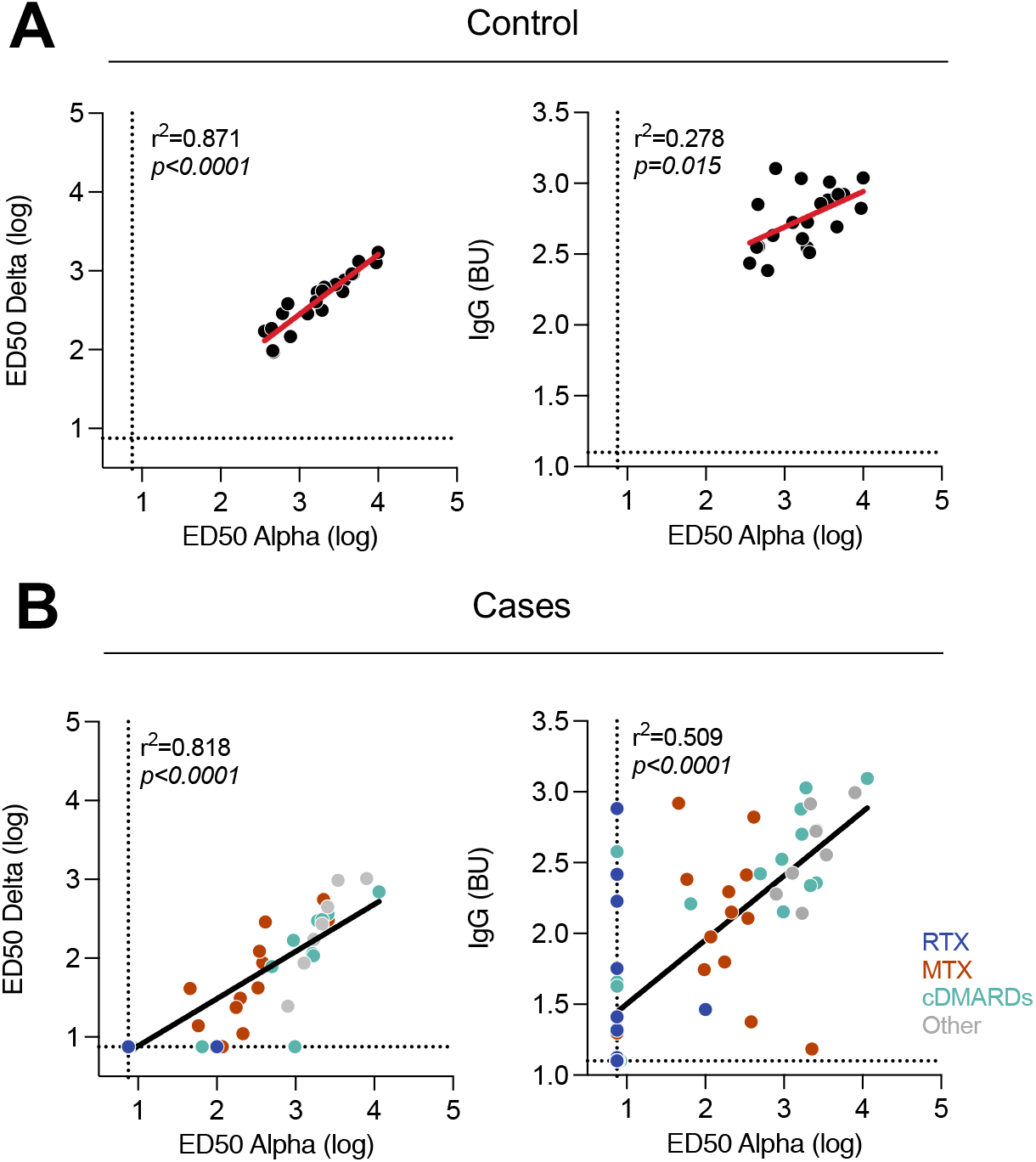
Cross-Neutralization Between Alpha and Delta Variants and Correlation with specific Anti-Spike IgG Levels at 3 Months.

**Figure S8.**
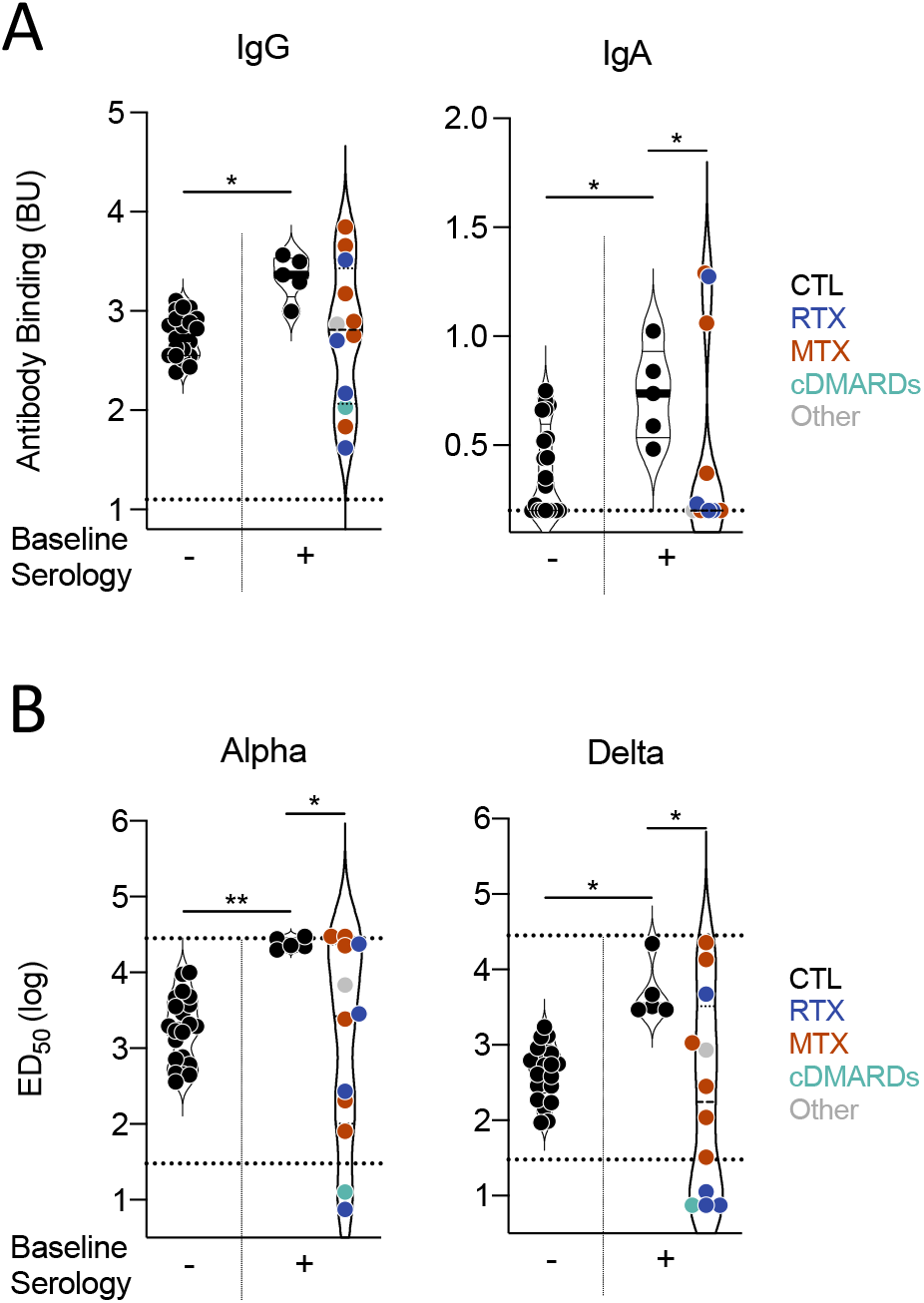
Humoral Immune Response to SARS-CoV-2 in Convalescent Vaccinated Individuals.

**Figure S9.**
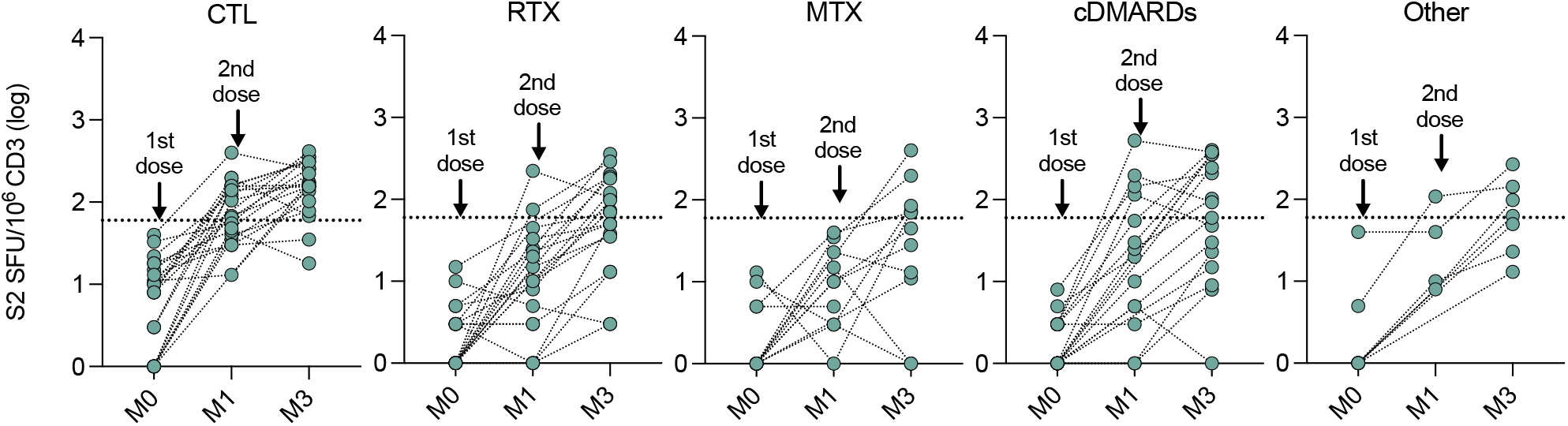
Kinetics of specific T-cell response against the SARS-CoV-2 S2 peptide pool.

