## Supplementary appendix for "Immunogenicity of BNT162b2 vaccine Against the Alpha and Delta Variants in Immunocompromised Patients": Supplementary appendix.pdf

#### Methods

##### *Patients*

The prospective COVADIS study (NCT04870411) was conducted in patients with systemic inflammatory diseases managed in the Internal Medicine department from Cochin Hospital, University of Paris (Paris, France). Healthcare immunocompetent workers from the same hospital were included as controls. Patients with a positive Covid-19 serology at baseline (day 0) were excluded from the main analysis. Cases and controls received the BNT162b2 mRNA vaccine from BioNTech/Pfizer according to the recommendations of the French National Authority for Health, considering either their immunocompromised status or their occupational exposure risk. Ethics approval was obtained by Comité de Protection des Personnes Nord-Ouest II. Cases and controls provided written informed consent.

##### *Clinical and laboratory data*

Clinical data were collected at baseline and during follow-up until month 3, and included demographics, underlying disease, disease activity, renal involvement, current and previous (within the 12 months before the first dose of vaccine) therapies. Total lymphocytes count and immunoglobulins dosage in peripheral blood were also collected at baseline. To evaluate vaccine immunogenicity, blood samples were collected before the first dose of vaccine (M0), before the second dose (M1) and 3 months after the first dose (M3).

##### *T and B cell immunophenotyping*

Briefly, after 2 washes with PBS 1X, FcR Blocking Reagent (Miltenyi) was used according to manufacturer instructions. Cellular stainings were performed at 4°C for 30 min protected from light. Antibodies used are summarized in the **Supplementary Table 2**. Samples were acquired using the BD LSRFortessa™ X-20 Cell Analyzer. Data were analyzed using FlowJo software v10.6. Gating strategies for immunoprofiling of B cell and T cell subpopulation compositions are shown in **Supplementary Figures 1-2**.

***S-Flow assay***

The S-Flow assay was adapted using 293T cells stably expressing the S protein (293T Spike cells) and 293T empty cells as control[1]. Cells were incubated at 4°C for 30 min with sera (1:300 dilution) in PBS containing 0.5% BSA and 2 mM EDTA. Cells were then washed with PBS, and stained using an anti-human IgG Fc Alexafluor 647 antibody (109-605-170, Jackson ImmunoResearch) and an anti-human IgA Alpha chain Alexafluor 488 antibody (109-545-011, Jackson ImmunoResearch). After 30min at 4°C, cells were washed with PBS and fixed for 10 min using 4% paraformaldehyde (PFA). Binding units (BU) were calculated to standardize the results. A standard curve with serial dilutions of a human anti-S monoclonal antibody (mAb48) was acquired in each assay. The logarithm of the median of fluorescence of each sample was reported on the curve to obtain an equivalent value (in ng/mL) of mAb48 concentration in logarithm [1,2]. Data were acquired on an Attune NxT instrument (Life Technologies). Flow cytometry data were analyzed with FlowJo v.10 software (TriStar). Calculations were performed using Excel 365 (Microsoft).

***Virus strains***

The B.1.1.7 (Alpha) variant originated from an individual in Tours (France) returning from the United Kingdom. The B.1.617.2 (Delta) variant originated from a hospitalized patient in Paris returning from India[3]. Both patients provided informed consent for the use of the biological materials. The variant strains were isolated from nasal swabs using Vero E6 cells and amplified by two passages. Titration of viral stocks was performed on Vero E6 cells, with a limiting dilution technique allowing a calculation of the 50% tissue culture infectious dose, or on S-Fuse cells. Viruses were sequenced directly on nasal swabs and after two passages on Vero cells.

***S-Fuse neutralization assay***

U2OS-ACE2 GFP1–10 or GFP 11 cells, also termed S-Fuse cells, were used, these cells becoming GFP+ cells when productively infected with SARS-CoV-2[3]. Cells were mixed (at a 1:1 ratio) and plated at $8.10^3$ cells per well in μClear 96-well plates (Greiner Bio-One). SARS-CoV-2 strains were incubated with sera at the indicated concentrations for 15 min at room temperature and added to S-Fuse cells. 18 h later, cells were fixed with 2% paraformaldehyde, washed and stained with Hoechst (1:1,000 dilution; Invitrogen). Images were acquired with an Opera Phenix high-content confocal microscope (PerkinElmer). The number of syncytia and nuclei were quantified using the Harmony software (PerkinElmer). The percentage of neutralization was calculated using the number of syncytia and the following formula: 100 x (1 − (value with serum – value in 'noninfected')/(value in 'no serum' − value in 'noninfected')). Neutralizing activity of each serum was expressed as the half maximal effective dilution (ED50), calculated using a reconstructed curve of neutralization at each concentration. Sera were heat-inactivated for 30 min at 56°C before use.

***T-cell response using enzyme-linked immunoSpot (EliSpot)***

Peripheral blood mononuclear cells (PBMCs) were isolated from fresh blood. After density gradient separation, lymphocytes were enumerated by flow cytometry using the BD Tritest™ CD3FITC/ CD8PE/ CD45PerCP and BD Trucount™ Tubes (BD Biosciences, Le Pont de Claix, France). The EliSpot assay was conducted as previously described[4,5]. Briefly, day 0, sterile PVDF strips (Millipore, Saint-Quentin-en-Yvelines, France) were coated overnight at 4°C with an IFN-γ antibody (U-CyTech, Utrecht, Netherlands). Day 1, strips were blocked with culture medium 1 hour and then, PBMCs were seeded at 200 000 CD3+T cell/well in duplicates and stimulated for 18-20hrs with individual pool of 15-mer peptides with 11 amino acids overlap at a final concentration of 10 μmol/L. Day 2, PBMCs were removed and IFN-γ secretion was revealed using a biotin-conjugated IFN-γ antibody (U-CyTech), streptavidinhorseradish peroxidase (UCyTech) and 3-amino-9-ethylcarbazole (AEC) (U-CyTech). Spots were enumerated using an automated EliSpot reader (Autoimmune Diagnostika (AID) reader, Strassberg, Germany). To identify SARS-CoV-2-spike-specific T cells, we used a commercially available pool derived from a peptide scan through SARS-CoV-2 N-terminal fragment (pool S1) (JPT Peptide Technologies GmbH, BioNTech AG, Berlin, Germany). Results were expressed as Spot Forming Unit (SFU)/106 CD3+ T lymphocytes after subtraction of background values from wells with non-stimulated cells. The EliSpot was technically validated when the mean number of spots in unstimulated wells was under than or equal to 10. The detection threshold was set at 3SD above the average basal reactivity (mean spot number in RPMI wells), fixed at a minimum of 11 SFU/$10^6$ CD3+ to rule out false positives where the background was very low.

***Statistical analysis***

No statistical methods were used to predetermine sample size. The experiments were performed in blind regarding to the allocation groups. Flow cytometry data were analyzed with FlowJo v.10 software (TriStar). Calculations were performed using Excel 365 (Microsoft). Figures were drawn using GraphPad Prism 9. Statistical analyses were conducted using GraphPad Prism 9. Statistical significance between different groups was calculated using the tests indicated in each figure legend. To assess clinical data and treatment associated with cross-neutralization of Alpha and Delta variants, and T-cell response at 3 months, patients were compared using *t*-tests for quantitative variables and analyses of variance for qualitative variables. Associations between both quantitative humoral and T-cell response (defined by neutralization titers for both viruses, and the number of circulating S1 and S2 peptide pool SARS-CoV-2 specific IFNγ-producing T cells) and clinical data were assessed by multivariate linear regression models. Those analyses were performed using R version 3.6.1. (R Foundation for Statistical Computing, Vienna, Austria).

### Tables

### Table S1. Immunological features at baseline

| Median (IQR) | Control n= 17 | Rituximab n=22 | Methotrexate n=16 | cDMARDs n=19 | Others n=7 |
|---|---|---|---|---|---|
| **Lymphocytes (/mm$^3$)** | 2400 (2125-2575) | 1100 (900-1780) | 1200 (110-2200) | 1570 (1100-1930 | 1600 (848-2125) |
| **CD19+ B cells (/mm3)** | 166 (105-210) | 0.1 (0-0.3) | 63 (49-203) | 180 (68-355) | 88 (29-141) |
| Naive B cells (%) | 67 (40-78) | - | 47 (18-70) | 73 (49-91) | 52 (51-74) |
| Switch memory B cells (%) | 14 (8-25) | - | 23 (11-33) | 7 (2-27) | 18 (15-31) |
| **CD4+ T cells (/mm3)** | 61 (56-65) | 51 (42-72) | 54 (47-67) | 58 (50-74) | 55 (41-69) |
| Naïve CD4+ T cells (%) | 41 (31-55) | 38.5 (19-53) | 31 (13-61) | 46 (35-56) | 24 (17-46) |
| Memory C4+ T cells (%) | 52 (39-61) | 55 (37-74) | 65 (33-82) | 48 (39-58) | 71 (50-80) |
| Follicular helper CD4+ T cells (%) | 8 (6-10) | 7 (5-9) | 7 (5-8) | 8 (6-13) | 9 (6-12) |
| **CD8+ T cells (%)** | 33 (28-36) | 37 (23-46) | 39 (29-41) | 31 (23-41) | 34 (25-46) |
| **Immunoglobulins, g/L** | | | | | |
| IgG | 11.1 (9.9-12.4) | 7 (6-8.6) | 10.4 (7.9-13.3) | 12.4 (8.6-18) | 11.6 (10.2-12.9) |
| IgA | 2.2 (1.5-2.6) | 1.4 (0.9-2) | 1.9 (1.5-2.8) | 1.7 (1.4-2.5) | 2.1 (1.8-3) |
| IgM | 0.9 (0.7-1.3) | 0.4 (0.2-0.8) | 0.9 (0.7-1.3) | 1 (0.5-1.3) | 0.8 (0.6-2.5) |
| IgG1 | 5.2 (4.2-6.2) | 3.8 (2.8-4.4) | 5.9 (3.6-7.3) | 5.4 (4.2-10.5) | 5.3 (4.6-6.1) |
| IgG2 | 3.5 (2.4-4.7) | 1.6 (1.3-2.1) | 2.4 (1.6-3.4) | 3.4 (2.2-4.2) | 3.4 (1.8-3.9) |
| IgG3 | 0.5 (0.3-0.6) | 0.3 (0.2-0.5) | 0.5 (0.3-0.7) | 0.8 (0.4-1.3) | 0.6 (0.3-0.7) |
| IgG4 | 0.55 (0.3-0.7) | 0.2 (0.1-0.4) | 0.15 (0.03-0.3) | 0.3 (0.1-0.4) | 0.3 (0.1-0.6) |

**Table S2. Antibodies used for T and B cell immunophenotyping.**

| | Marker | Fluorochrome | Clone | Supplier | Cat. number |
|---|---|---|---|---|---|
| Panel B | CD19 | APC | SJ25C1 | Biolegend | 363006 |
| | IgD | PE | I-A62 | BD Pharmigen | 555779 |
| | CD21 | FITC | BL13 | Beckman Coulter | IMO473U |
| | CD27 | BV510 | O323 | Sony | 2114180 |
| | CD24 | BV421 | ML5 | BD Horizon | 562789 |
| | CD38 | PE-Cy7 | HB-7 | Biolegend | 356608 |
| Panel T | CD3 | PerCP-Cy5.5 | OKT3 | Biolegend | 317336 |
| | CD4 | BV510 | OKT4 | Biolegend | 317444 |
| | CD8 | APC | HiT8a | Biolegend | 300912 |
| | CD45RA | AF488 | Hi100 | Biolegend | 304114 |
| | CD38 | PE-Cy7 | HB-7 | Biolegend | 356608 |
| | CCR6 | BV650 | G034E3 | Biolegend | 353426 |
| | CXCR5 | BV711 | J252D4 | Sony | 2384665 |
| | PD-1 | PE | EH12.2H7 | Biolegend | 329906 |
| | CXCR3 | PE-Dazzle | G025H7 | Biolegend | 353736 |
| | ICOS | AF700 | C398.4A | Sony | 2167640 |

**Table S3. Characteristics of patients according to the neutralization of alpha and delta variants and to the T-cell response (n=82).**

| | Neutralization of alpha variant | | | Neutralization of delta variant | | | T-cell response | | |
|---|---|---|---|---|---|---|---|---|---|
| | No | Yes | *P* | No | Yes | *P* | No | Yes | *P* |
| N | 30 | 52 | | 39 | 43 | | | | |
| Age, years | 55.47 (16.64) | 50.13 (15.42) | 0.147 | 2.56 (17.26) | 51.65 (14.94) | 0.798 | 48.05 (17.90) | 53.96 (15.31) | 0.169 |
| Male | 9 (30.0) | 14 (26.9) | 0.965 | 23 (28.0) | 11 (28.2) | 1.000 | 5 (26.3) | 15 (27.3) | 1.000 |
| Treatment group | | | <0.001 | | | <0.001 | | | 0.036 |
| Controls | 0 (0.0) | 21 (40.4) | | 0 (0.0) | 21 (48.8) | | 1 (5.3) | 18 (32.7) | |
| cDMARD | 9 (30.0) | 10 (19.2) | | 11 (28.2) | 8 (18.6) | | 6 (31.6) | 11 (20.0) | |
| Methotrexate | 2 (6.7) | 13 (25.0) | | 7 (17.0) | 8 (18.6) | | 6 (31.6) | 5 (9.1) | |
| Rituximab | 19 (63.3) | 1 (1.9) | | 20 (51.3) | 0 (0.0) | | 5 (26.3) | 15 (27.3) | |
| Other | 0 (0.0) | 7 (13.5) | | 1 (2.6) | 6 (14.0) | | 1 (5.3) | 6 (10.9) | |
| Group of disease | | | <0.001 | | | <0.001 | | | 0.115 |
| Controls | 0 (0.0) | 21 (40.4) | | 0 0.0) | 21 (48.8) | | 1 (5.3) | 18 (32.7) | |
| CTD | 9 (30.0) | 20 (38.5) | | 14 (35.0) | 15 (34.9) | | 9 (47.4) | 17 (30.9) | |
| Vasculitis | 20 (66.7) | 5 (9.6) | | 22 (56.4) | 3 (7.0) | | 7 (36.8) | 17 (30.9) | |
| Other | 1 (3.3) | 6 (115) | | 3 (7.7) | 4 (9.3) | | 2 (10.5) | 3 (5.5) | |
| Active disease (N=61) | 9 (30.0) | 7 (22.6) | 0.713 | 12 (30.8) | 4 (9.3) | <0.001 | 7 (38.9) | 6 (16.2) | 0.129 |
| Glucocorticoids (%) | 21 (70.0) | 22 (42.3) | 0.029 | 27 (69.2) | 16 (37.2) | 0.007 | 15 (78.9) | 23 (41.8) | 0.012 |
| Lymphocytes, *per mm3* | 1340.36 (725.59) | 1618.36 (690.38) | 0.126 | 1434.59 (745.91) | 1580.48 (671.36) | 0.429 | 1418.89 (731.18) | 1544.68 (714.65) | 0.542 |
| IgG, g/L | 9.34 (5.70) | 12.41 (5.81) | 0.025 | 10.74 (7.47) | 11.72 (3.85) | 0.470 | 13.18 (10.41) | 10.45 (3.22) | 0.094 |
| IgA, g/L | 1.46 (0.67) | 2.38 (1.50) | 0.002 | 1.82 (1.60) | 2.23 (0.93) | 0.175 | 2.02 (1.25) | 2.10 (1.42) | 0.830 |
| IgM, g/L | 0.69 (0.61) | 1.06 (0.61) | 0.012 | 0.78 (0.64) | 1.06 (0.60) | 0.047 | 0.89 (0.61) | 0.90 (0.66) | 0.950 |
| IgG1, g/L | 5.11 (3.64) | 6.12 (3.21) | 0.204 | 5.74 (4.13) | 5.73 (2.50) | 0.990 | 7.05 (5.75) | 5.15 (1.82) | 0.037 |
| IgG2, g/L | 2.05 (1.13) | 3.53 (1.78) | <0.001 | 2.58 (1.98) | 3.34 (1.31) | 0.051 | 2.95 (2.53) | 3.00 (1.36) | 0.915 |
| IgG3, g/L | 0.56 (0.59) | 0.58 (0.28) | 0.814 | 0.63 (0.56) | 0.52 (0.21) | 0.263 | 0.70 (0.65) | 0.50 (0.23) | 0.065 |
| IgG4, g/L | 0.21 (0.18) | 0.36 (0.30) | 0.019 | 0.21 (0.18) | 0.39 (0.32) | 0.003 | 0.20 (0.21) | 0.33 (0.30) | 0.098 |

Data are presented as mean (SD) for continuous variables and as count (percent) for qualitative variables. Comparisons between patients with humoral or cellular response were compared using t-tests for continuous variables and analyses of variance for qualitative variables.

**Table S4. Multivariate linear regression models assessing the association between patient's characteristics and quantitative humoral and cellular response.**

| | ED50 alpha | | ED50 delta | | SARS-CoV-2-specific IFNγ-producing T cells | |
|---|---|---|---|---|---|---|
| | β coefficient [95%CI] | *P* | β coefficient [95%CI] | *P* | β coefficient [95%CI] | *P* |
| Age, years | 7.49 [-25.17, 40.14] | 0.649 | 2.16 [-1.91, 6.22] | 0.294 | -1.61 [-4.41, 1.18] | 0.253 |
| Treatment group | | | | | | |
| Controls | Ref | | Ref | | Ref | Ref |
| cDMARD | -1809.56 [-3590.36, -28.77] | 0.047 | -434.85 [-669.67, -200.03] | <0.001 | 26.40 [-126.56, 179.35] | 0.731 |
| Methotrexate | -2729.50 [-4485.78, -973.23] | 0.003 | -462.83 [-701.35, -224.31] | <0.001 | -70.95 [-227.87, 85.97] | 0.370 |
| Rituximab | -3153.98 [-4823.90, -1484.06] | <0.001 | -583.41 [-803.88, -362.95] | <0.001 | 77.73 [-62.81, 218.26] | 0.273 |
| Other | -398.73 [-2351.02, 1553.55] | 0.685 | -190.32 [-440.08, 59.43] | 0.133 | -35.00 [-198.04, 128.04] | 0.669 |
| Glucocorticoids (%) | -50.01 [-1332.51, 1232.50] | 0.938 | -48.87 [-207.06, 109.31] | 0.540 | -44.34 [-153.91, 65.23] | 0.4222 |
| IgA, g/L | 31.61 [-279.05, 342.27] | 0.840 | 0.10 [-41.14, 41.34] | 0.996 | 45.96 [14.67, 77.25] | 0.005 |
| IgG2, g/L | 189.43 [-183.08, 561.95] | 0.314 | 246.59 [-29.18, 522.36] | 0.079 | -11.57 [-38.53, 15.38] | 0.394 |

### Figure legends

**Figure S1. Gating Strategy for Immunoprofiling of B cell Subpopulation Compositions.**

Lymphocytes were first gated based on their FSC-A versus SSC-A. CD19+ B cells were separated between naive (CD27-) and memory (CD27+) subsets then IgD marker helped defined switched memory (CD27+IgD-), marginal zone (CD27+IgD+) and naive (CD27-IgD+) CD19+ B cells. Plasmablasts were defined as CD24-CD38high among memory CD19+ B cells.

**Figure S2. Gating Strategy for Immunoprofiling of T cell Subpopulation Compositions.**

Lymphocytes were first gated based on their FSC-A versus SSC-A. Within the CD3+ T cell compartment, CD4+ T cells were further subdivided into naïve (CD45RA+) and memory (CD45RA-). Among memory CD4+ T cells, PD-1 and CXCR5 surface markers were used to define follicular helper T cells (TFH, CXCR5+PD-1+CD45RA-CD4+). Identification of Th1 (CXCR3+CXCR6-), Th2 (CXCR3-CXCR6-), Th17 (CXCR3-CXCR6+) and Th1+Th17+ double positive (CXCR3+CXCR6+) populations were identified among CXCR5-CD45RA-CD4+ subset.

**Figure S3. Laboratory Findings in Patients Included in the Study.**

**A.** Lymphocyte counts, absolute number of CD3+ T cells, CD19+ B cells and proportions (frequencies) of CD19+ B cells, CD8+ T cells and CD4+ T cells among lymphocytes in peripheral blood according to the treatments received.

**B**. Concentration of immunoglobulin G, immunoglobulin A, immunoglobulin M proteins measured in serum. Data indicate median. Each dot represents a single patient. Two-sided Kruskal-Wallis test with Dunn's test for multiple comparisons between group of treatment was performed with median reported; *P < 0.05; **P < 0.01; ***P < 0.001.

**Figure S4. Immunoprofiling of Peripheral Blood Mononuclear Cells at Baseline.**

Proportions (frequencies) in peripheral blood of naive B cells and switched memory B cells among CD19+ B cells, naïve CD4+ T cells and memory CD4+ T cells among CD4+ T cells, follicular helper CD4+ T cells and TH1 CD4+ T cells among CD4+ memory CD4+ T cells and CD8+ T cells among lymphocytes according to the treatments received. Data indicate median. Each dot represents a single patient.

**Figure S5. Kinetics of Anti-spike IgG and IgA after BNT162b2 Vaccine.**

**A-B.** Levels of anti-S IgG **(A)** and IgA **(B)** antibodies (defined as binding units; BU) before the first dose of vaccine (M0), before the second dose (M1) and after full vaccination at 3 months (M3) according to the treatments received. The dotted line indicates threshold (BU=1.1 for IgG and 0.2 for IgA).

**Figure S6. Kinetics of TH1 CD4+ T cells, Follicular Helper CD4+ T cells, Plasmablasts and Switched Memory B cells After BNT162b2 Vaccine.**

Proportions (frequencies) in peripheral blood of switched memory B cells among CD19+ B cells and follicular helper CD4+ T cells among memory CD4+ T cells before the first dose (M0), before the second dose (M1) and after full vaccination at 3 months (M3) according to the treatments received.

**Figure S7. Cross-Neutralization of the Alpha and Delta Variants and Correlation with specific Anti-Spike IgG Levels at 3 Months.**

**A-B.** Correlation between Alpha and Delta neutralization titers (ED50) (left panel) or between IgG levels and Alpha neutralization titer (right panel). **(A)** control group and **(B)** patient groups. A Spearman correlation model was applied. p and $r^2$ values are indicated.

**Figure S8. Humoral Immune Response to SARS-CoV-2 in Convalescent Vaccinated Individuals.**

**A.** Level of anti-S IgG and IgA antibodies at M3 post-vaccination in the control group (black dots) and cases groups (color dots) with a positive (+) or negative (-) serology before vaccination. The dotted line indicates threshold (BU=1.1 for IgG and 0.2 for IgA).

**B.** Neutralizing titers of sera against Alpha (left panel) and Delta (right panel) variants are expressed as ED50 values. The lower limit of detection (ED50=30) and the upper line of quantification (ED50=30,000) are indicated by dotted lines.

Two-sided Kruskal-Wallis test with Dunn's test for multiple comparisons was performed. *P<0.05, **P<0.01, ***P<0.001, ****P<0.0001.

**Figure S9. Kinetics of specific T-cell response against the SARS-CoV-2 S2 peptide pool.**

Kinetic of specific T-cells responses against the SARS-CoV-2 S2 peptide before the first dose of vaccine (M0), before the second dose (M1) and after full vaccination at 3 months (M3) according to the treatments received. Data indicate median. Each dot represents a single patient.
